# Survey of Behaviour Attitudes Towards Preventive Measures Following COVID-19 Vaccination

**DOI:** 10.1101/2021.04.12.21255304

**Authors:** Daniella Rahamim-Cohen, Sivan Gazit, Galit Perez, Barak Nada, Shay Ben Moshe, Miri Mizrahi-Reuveni, Joseph Azuri, Tal Patalon

## Abstract

Following the widespread vaccination program for COVID-19 carried out in Israel, a survey was conducted to preliminarily assess behavior changes in the vaccinated population, prior to the expected upcoming policy change as to mask wearing and social distancing regulation in Israel. 200 people answered at least one question pertaining to preventive behaviour. Among the respondents, 21.1% reported a decrease in mask wearing compared to 47.3% who reported a decrease in social distancing. There was no difference in these measures between the sexes. However, people under the age of 50 were more likely to decrease mask wearing (28.1%) and decrease social distancing (56.1%), as compared with people over the age of 50 (17.2% and 41.8%, respectively). Among health care workers, there was a minimal decrease in mask wearing (1/23 people) compared to a more widespread decrease in social distancing (10/23). These data suggest that preventive attitudes change following COVID-19 vaccination, with less adherence to social distancing as compared to mask wearing, and should be taken into account when planning public policy in the future.

## Introduction

Since the onset of the COVID-19 global pandemic in late 2019, there have been over 130,000 million reported cases with over 3 million deaths worldwide. Most countries set various guidelines and procedures in an attempt to curb the spread of the virus, including the use of personal protective equipment (PPE) and maintaining social distancing. These measures were found to be very effective in reducing viral transmission rate^1^ and were the cornerstones for health policy in tackling virus spread in most countries around the world, manifesting in lockdowns and various enforcement measures to ensure compliance.

With the introduction of vaccines against the virus, the question arises about a possible change in personal attitudes and behaviours to the recommendations for mask wearing and keeping social distancing.

Evidence from previous vaccine rollouts suggests that there may be a decrease in adherence to preventive behaviours. Following the Lyme disease vaccine rollout, there was a decrease in the use of light color clothing and tick repellent^2^. An evaluation of behvaiour following the influenza vaccine’s rollout showed that people interacted with more people and in larger groups^3^.

Israel began vaccinating its population at the end of December 2020, using the Pfizer/BioNtech BNT162b2 COVID-19 vaccine. The vaccination program was implemented through HMO’s and hospitals, with clear prioritization – beginning with the elderly and medical staff, until the current situation where any citizen over the age of 16 is eligible to receive a vaccine.

As of April 8^th^, 52.8% of the Israeli population has completed the course of two vaccinations^4^ with over 5 million people vaccinated. Since the beginning of the program, Israel has seen a steady decrease in confirmed COVID-19 cases and mortality. As a result of the improvement in morbidity and mortality and the high vaccination uptake, the government has lifted many of the restrictions imposed on the population over the past year, opening shops, restaurants, and schools. Some are accessible only to fully vaccinated individuals. However, the policy to wear masks indoors and outdoors and maintain social distancing is still in effect and has not officially changed.

The possible effect of the COVID-19 vaccination program on adherence to the rules and regulations surrounding personal protective behaviour is central to formulating health policy regarding the virus in the vaccination and post-vaccination era. The UK has prepared an executive summary regarding the possible impact of widespread vaccination on adherence outcomes and how they can be mitigated^5^. To assess Israel’s situation and explore if there are behavioural changes in the population following vaccination, we conducted a survey to identify behaviour attitudes in the general vaccinated population within Maccabi Healthcare Services (MHS).

## Method

A survey was sent by a text message to 2500 subjects, randomly seleted, above the age of 18, vaccinated or not vaccinated, from MHS, the second largest HMO in Israel, serving approximately 2.5 million people (about a quarter of the Israeli population) with nationwide distribution. The survey was conducted between 25/3/21-7/4/21. Participants were asked to compare mask-wearing and social distancing before and after the vaccination. Demographics included age, sex, and being a healthcare professional.

The following analysis is preliminary, as we are currently conducting further research exploring the influence of the upcoming healthcare policy changes.

The study was approved by the MHS institutional review board.

## Results

Of 2500 surveys sent, 200 people aged 18-80 years responded to at least one question. The average time for answering was 3:39 minutes. Thirteen respondents were not vaccinated with even one vaccination. The main results are described in Table 1.

**Table 1.** Main results.

|  |  | All | Male | Female | Age <50 | Age ≥50 | Health workers |
| --- | --- | --- | --- | --- | --- | --- | --- |
| Mask usage post vaccination | Responded | 185 | 80 | 105 | 57 | 122 | 23 |
|  | Lower | 39 (21.1%) | 16 (20.5%) | 23 (21.9%) | 16 (28.1%) | 21 (17.2%) | 1 (4.3%) |
|  | No change | 140 (75.7%) | 62 (77.5%) | 78 (74.3%) | 40 (70.2%) | 97 (79.5%) | 22 (95.7%) |
|  | Higher | 6 (3.2%) | 2 (2.5%) | 4 (3.8%) | 1 (1.8%) | 4 (3.3%) | 0 (0%) |
| Social distancing post vaccination | Responded | 184 | 80 | 104 | 57 | 122 | 23 |
|  | Lower | 87 (47.3%) | 38 (47.5%) | 49 (47.1%) | 32 (56.1%) | 51 (41.8%) | 10 (43.5%) |
|  | No change | 89 (48.4%) | 39 (48.8%) | 50 (48.1%) | 23 (40.4%) | 65 (53.3%) | 12 (52.2%) |
|  | Higher | 8 (4.3%) | 3 (3.8%) | 5 (4.8%) | 2 (3.5%) | 6 (4.9%) | 1 (4.3%) |

Following the second vaccination, 39 respondents reported “lower mask wearing” than before receiving the vaccination (21.1%) compared to 140 who did not change their mask wearing behaviour (75.7%) [Figure 1]. There was no substantial difference in terms of sex or being a healthcare professional in terms of decreased mask wearing; however, more people aged less than 50 decreased their mask wearing, 28.1% (16/57) compared to the group aged 50 and above, 17.2% (21/122).

**Figure 1.**
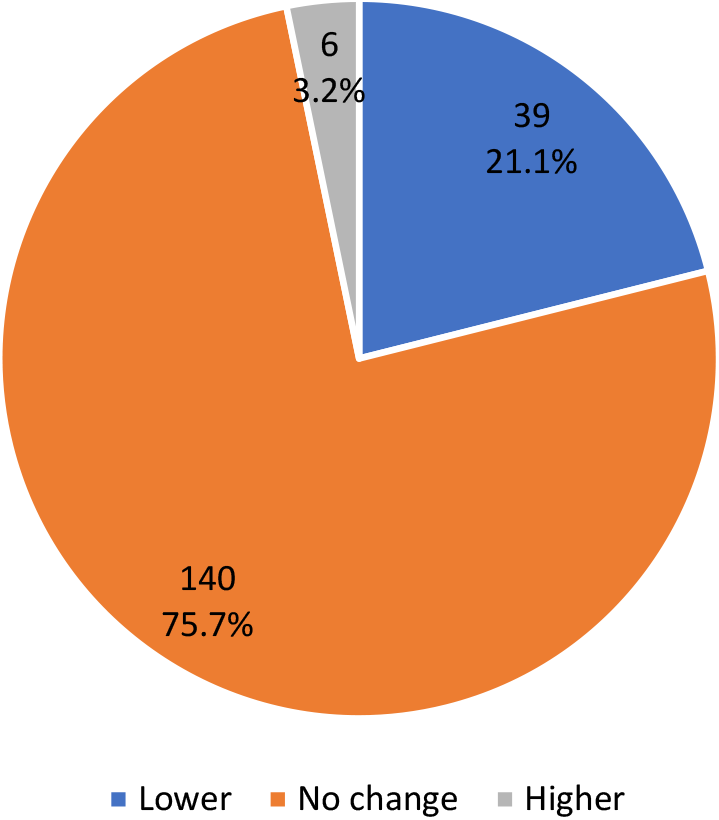
Mask wearing behaviour post vaccination

When comparing social distancing behavior before and after vaccination, 89 (48.4%) reported no change, whereas 87 (47.3%) respondents reported a decrease in social distancing [Figure 2]. There was only a slight difference between males and females (38/80, 47.5%; 49/104, 47.1%, respectively).

**Figure 2.**
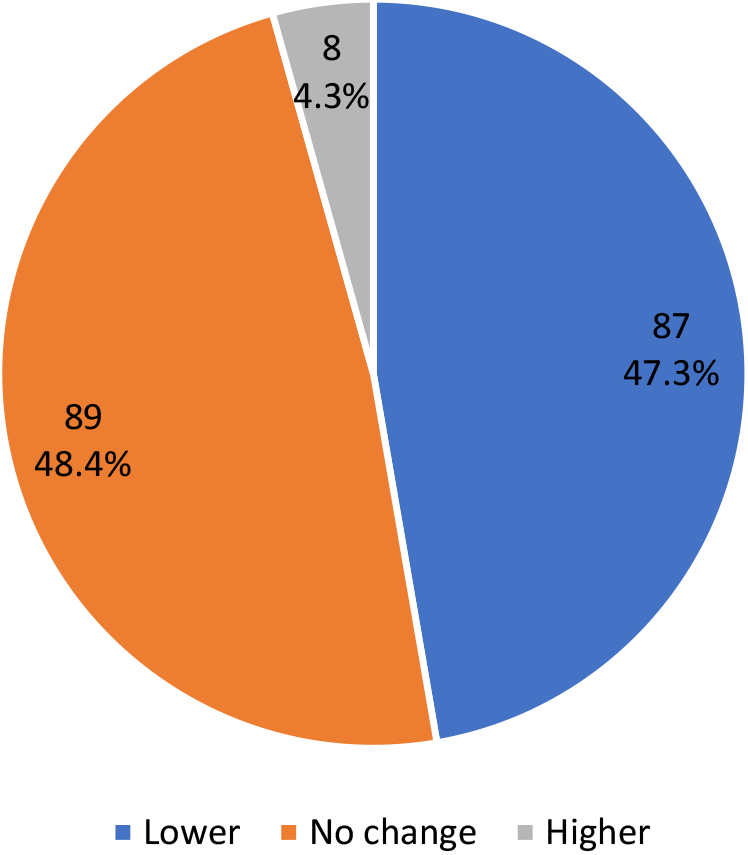
Social distancing practices post vaccination

Of the respondents aged 50 and above, 41.8% (51/122) reported keeping “less social distancing”, while 53.3% (65/122) reported no change, whereas respondents below the age of 50 reported 56.1% (32/57) and 40.4% (23/57) respectively.

Only one healthcare professional reported decreased “mask wearing” following the vaccine (1/23), in comparison to almost half (10/23) of the healthcare professionals who reported a decrease in “social distancing” following the vaccine.

## Discussion

Consistent with previous findings^2,3^, the survey showed decreased social distancing and mask wearing in specific population groups following COVID-19 vaccination. The reported decrease in mask wearing was less prevalent than the reported decrease in social distancing, which occurred in approximately half the surveyed population. In the subgroup of health professionals, it is clear that maintaining mask-wearing is still the prevailing practice, as opposed to maintaining social distancing.

Moreover, the survey suggests that people under the age of 50 are less likely to maintain the preventive behaviours of mask wearing and social distancing when compared to the older population.

Although the data above is not surprising, it should be taken into account when devising health policy regarding COVID-19 preventive behaviour in populations that have been widely vaccinated.

## Limitations

The current survey is a preliminary effort to assess health behaviour in vaccinated population, prior to the upcoming policy change as to mask wearing and social distancing regulation in Israel. The cohort is relatively small, without statistical analysis. Availability of cell phone use among different populations (e.g. ultraorthodox, elderly) is of consideration, as further studies will address these issues.

## Data Availability

Data is part of the Maccabi Healthcare Services trial database and accessible upon request.

